# Vaccine uptake patterns for COVID-19 and cholera among healthcare workers: a cross-sectional study in Yaoundé-Cameroon

**DOI:** 10.64898/2026.03.12.26348275

**Authors:** Ariane Nouko, Fabrice Zobel Lekeumo Cheuyem, Félicitée Nguefack, Rick Tchamani, Innocent Takougang

## Abstract

**Objective:** Healthcare workers (HCWs) are at increased risk of COVID-19 infection and play a critical role in influencing public vaccine acceptance. This study aimed to assess vaccination coverage and identify the determinants of vaccine uptake among healthcare workers in Cameroon, in order to inform targeted strategies to strengthen immunization programs and improve epidemic preparedness.

**Results:** Among 406 participants (75.6% female, 65.5% aged 30-44 years, 61.3% nurses), 42.4% were fully vaccinated against COVID-19, while only 4.7% had completed the cholera vaccine series. Coverage varied across districts, with Biyem-Assi (53.0%) and Odza (46.0%) recording the highest COVID-19 uptake, and Nkolndongo (11.5%) leading for cholera vaccination. Independent predictors of COVID-19 uptake included being a nurse (aOR = 3.96; 95% CI: 2.07-7.81; *p* = 0.001) and laboratory technician professions (aOR = 8.00; 95% CI: 1.38-69.8; *p* =0.032). For cholera vaccination, working in internal medicine wards (aOR=11.2; 95% CI: 1.04-120; *p* = 0.046) and being a nurse (aOR = 1.54; *p* = 0.001) emerged as significant predictors. Although 62.8% of HCWs perceived their work environment as high-risk, knowledge of recommended vaccines was limited, with only 18.7% aware of cholera vaccination recommendations. Strengthening vaccine education, improving accessibility, and reducing financial barriers could enhance vaccine acceptance among HCWs. These findings provide important insights for designing targeted immunization strategies in Cameroon and similar contexts.

## Introduction

Infectious diseases remain a major public health challenge worldwide, characterized by both an increasing incidence in some regions and the re-emergence of previously controlled conditions [1, 2]. Healthcare facilities represent high-risk environments due to the high concentration of pathogens and the frequency of contact between patients, visitors, and healthcare workers, thereby facilitating disease transmission [3]. Globally, the rate of hospital-acquired infections is estimated at approximately 0.14% [4]. In 2019, infectious diseases were responsible for an estimated 13.7 million deaths worldwide, highlighting their persistent impact on global morbidity and mortality [5]. In this context, healthcare workers play a central role in transmission dynamics, potentially serving as a source of pathogen spread to patients and their relatives, while also being themselves at increased risk of infection [6, 7].

Vaccines remain among the most effective public health interventions for reducing morbidity and mortality associated with infectious diseases [8, 9]. Evidence indicates that COVID-19 vaccination reduced deaths by 59% in Europe, corresponding to approximately 1.6 million lives saved [10]. In Cameroon, the national vaccination response was implemented following confirmation of the first COVID-19 case on March 6, 2020, with health authorities rapidly integrating vaccination into key preventive strategies [11]. Four vaccines were subsequently approved by national health regulatory bodies [12]. Despite these efforts, vaccine coverage remained relatively low, with only 6.8% of the eligible population vaccinated as of July 22, 2023 far below the global target of 70% set for the same period [13]. This limited uptake occurred within a context of substantial vaccine hesitancy driven by risk misperception, insufficient knowledge about vaccines, fear of adverse effects, and the widespread dissemination of misinformation [14]. In 2023 alone, COVID-19 caused 1,974 deaths in Cameroon, with a case-fatality rate estimated at 1.6%, underscoring the continuing burden of the disease [15].

Concurrently, Cameroon continues to experience recurrent cholera outbreaks, largely driven by environmental and structural determinants such as inadequate access to safe drinking water, poor urban sanitation, and seasonal rainfall patterns that facilitate disease transmission [16]. In 2023, cholera resulted in 478 deaths, corresponding to a case-fatality rate of 2.7% [17]. In this context, oral vaccination represents a critical preventive tool, particularly through the use of Euvichol and Shanchol vaccines, which have been widely deployed during national vaccination campaigns according to epidemiological needs and vaccine availability [18, 19]. Nevertheless, vaccine acceptance remains limited, with misinformation identified as a major barrier and refusal rates reaching up to 67% in certain localities [20].

Despite ongoing vaccination efforts, significant gaps remain in understanding the determinants of vaccine uptake in Cameroon, particularly among healthcare workers. Although they are prioritized for immunization and play a key role in influencing public trust, vaccine hesitancy persists within this group [21–23]. This study therefore, aimed to assess vaccination coverage and identify the determinants of vaccine uptake among healthcare workers in Cameroon, in order to inform targeted strategies to strengthen immunization programs and improve epidemic preparedness.

## Methods

### Study design and period

We conducted a hospital-based cross-sectional and analytical study in the seven (7) DHs in Yaoundé from January to June 2024.

### Study setting

Yaoundé is the political capital of the Centre region and of Cameroon. Its population was estimated at 4.85 million in 2025. It is the country’s second-largest city, and all ethnic groups in Cameroon are well represented in this city. The Cameroonian health system is organized around health districts, which are the operational level of healthcare implementation. A district hospital is the first level of reference in the health pyramid and provides primary healthcare. The seven DHs in Yaoundé, namely Biyem-Assi, Cité-Verte, Djoungolo-Olembe, Efoulan, Mvog-Ada, Nkolndongo, and Odza, cover a population of approximately 4.2 million residents, cumulatively employing nearly 560 health personnel, providing 650 beds, and handling 216,235 consultations and 25,320 admissions per annum.

### Participants

Our study population included all healthcare workers providing care to the general population in Yaoundé health facilities. Meanwhile, study participants encompassed HCWs in the following clinical units at each DH: surgery, internal medicine, obstetrics and gynaecology, laboratory, paediatrics, emergency, outpatient department, and hygiene department who have provided their written informed consent to participate in this study.

### Sampling method

The sample size was calculated using the single proportion formula (n=[Zα/2]2 *[P (1-P)] /E2) at a 95% confidence interval, where Zα / 2=1.96 and P=44.8%. Using a standard error of E=5%, a 10% dropout rate, a minimum sample size of 380 was obtained. In each clinical department, all consenting personnel were enrolled in the study.

### Data collection

Data was collected using an anonymous, structured and self-administered questionnaire, which consisted of 28 questions organized in four sections. It captured data related to socio-demographic characteristics, knowledge on vaccination, working environment, vaccination coverage, and compliance

### Variable and operational definition

Independent variables included sex, age group, professional experience in years, health unit in which HCWs worked, and the HCWs’ grade. The dependent variable was vaccination coverage for COVID-19 and Cholera vaccines. HCWs were considered as fully vaccinated if they had completed the primary series of COVID-19 vaccine (1dose Johnson and Johnson vaccine or 2 doses or more of Pfizer/AstraZeneca). HCWs who had completed at least 2 doses of oral Cholera vaccine (Shanchol or Euvichol) were considered fully vaccinated.

### Data processing and analysis

All filled questionnaires were cross-checked, entered, recoded as necessary, and analysed using R Statistics Version 4.3.3. Categorical variables were described using frequency (n) and percentage (%). The Fisher exact test was used to compare proportions. Simple and multiple binary logistic regressions were used to assess the strength of the association between variables and adjust for potential confounders. The predictors that best fit the model were chosen step by step using the Akaike Information Criterion (AIC). The model with the lowest index was selected. A *p*-value <0.05 was considered statistically significant. Confidence intervals (CIs) were estimated at a 95% level of confidence.

### Ethical approval statement

This study received authorization from the Faculty of Medicine and Biomedical Sciences IRB (N° 1115/UY1/FMSB/VDRC/DAASR/CSD) and from the Institutional Review Board of the Regional Delegation of Public Health for the Centre Region (N° 0244-3/CRERSHC/2024). Informed consent was obtained from all participants prior to their inclusion in the study. All methods were performed in accordance with the Declaration of Helsinki.

## Results

### Socio-professional characteristics of study participants

Out of the 450 HCWs contacted, 435 gave their consent, 406 returned the completed questionnaire, representing a 90% response rate. Most of our study participants were female (75.6%). Participants aged 30-45 years were the most represented (65.5%). They were mostly nurses (61.3%) and medical doctors (17.0%) (**Table 1**).

**Table 1.**
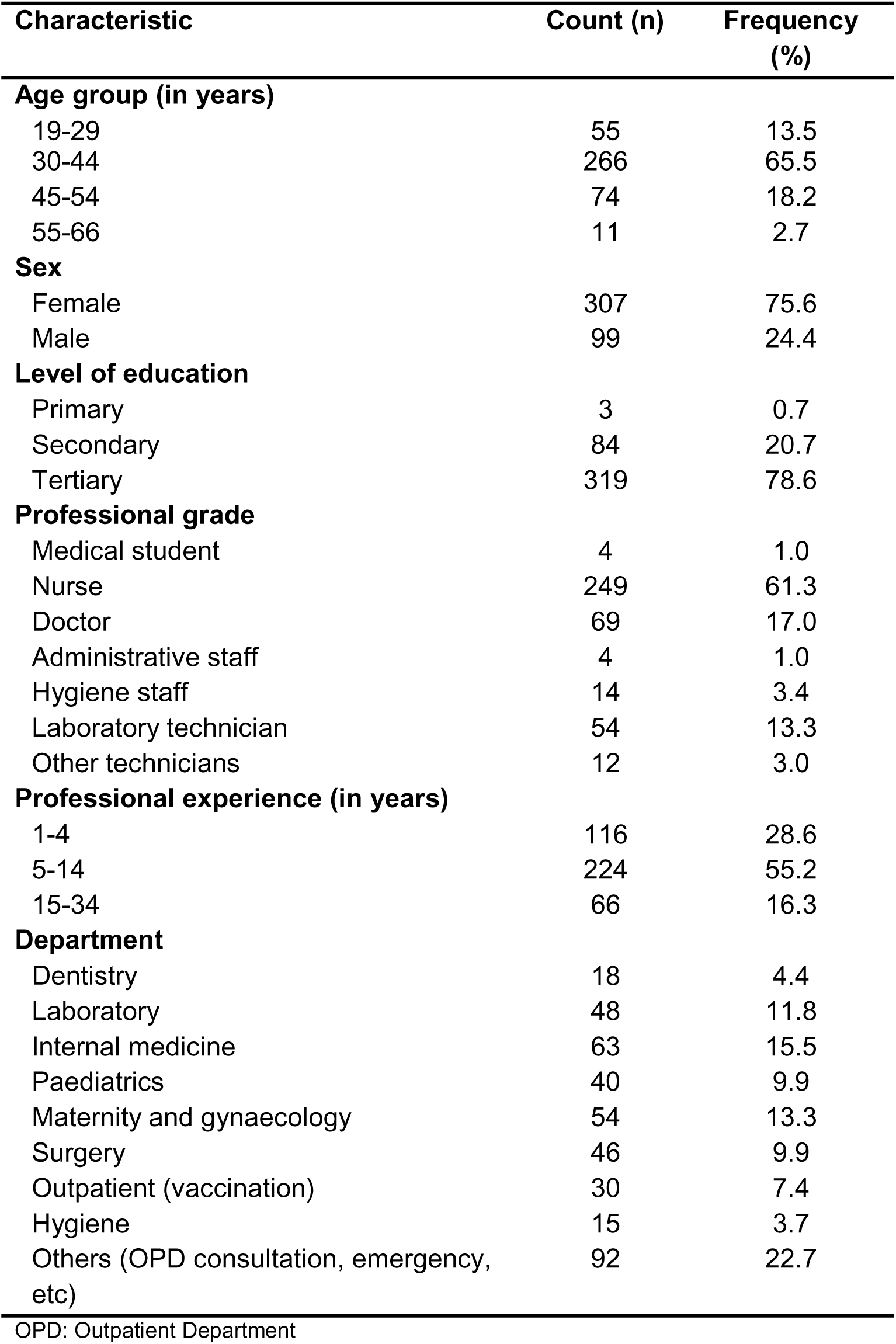
Socio-professional characteristics of study participants, Yaoundé District Hospitals, June 2024 (n = 406)

| Characteristic | Count (n) | Frequency (%) |
| --- | --- | --- |
| **Age group (in years)** |  |  |
| 19-29 | 55 | 13.5 |
| 30-44 | 266 | 65.5 |
| 45-54 | 74 | 18.2 |
| 55-66 | 11 | 2.7 |
| **Sex** |  |  |
| Female | 307 | 75.6 |
| Male | 99 | 24.4 |
| **Level of education** |  |  |
| Primary | 3 | 0.7 |
| Secondary | 84 | 20.7 |
| Tertiary | 319 | 78.6 |
| **Professional grade** |  |  |
| Medical student | 4 | 1.0 |
| Nurse | 249 | 61.3 |
| Doctor | 69 | 17.0 |
| Administrative staff | 4 | 1.0 |
| Hygiene staff | 14 | 3.4 |
| Laboratory technician | 54 | 13.3 |
| Other technicians | 12 | 3.0 |
| **Professional experience (in years)** |  |  |
| 1-4 | 116 | 28.6 |
| 5-14 | 224 | 55.2 |
| 15-34 | 66 | 16.3 |
| **Department** |  |  |
| Dentistry | 18 | 4.4 |
| Laboratory | 48 | 11.8 |
| Internal medicine | 63 | 15.5 |
| Paediatrics | 40 | 9.9 |
| Maternity and gynaecology | 54 | 13.3 |
| Surgery | 46 | 9.9 |
| Outpatient (vaccination) | 30 | 7.4 |
| Hygiene | 15 | 3.7 |
| Others (OPD consultation, emergency, etc) | 92 | 22.7 |

OPD: Outpatient Department

### Vaccination coverage for COVID-19 and Cholera

More than half of the HCWs in Yaoundé (57.4%) had not taken the COVID-19 vaccine, while 42.4% had taken full doses of the COVID-19 vaccine. Most of the HCWs had not taken the Cholera vaccine (80.3%), while a low proportion of HCWs had taken the complete dose of the Cholera vaccine (4.7%) (**Table 2**).

**Table 2.** Vaccination coverage among healthcare workers in Yaoundé District Hospitals, June 2024 (n=406)

| Type of vaccine | District hospital | Fully | Partially | Unvaccinated | Total (100%) | p-value* |
| --- | --- | --- | --- | --- | --- | --- |
| COVID-19 |  |  |  |  |  | 0.388 |
|  | Biyem-Assi | 35 (53.0) | 0 (0.0) | 31 (47.0) | 66 |  |
|  | Cite-Verte | 24 (39.3) | 0 (0.0) | 37 (60.7) | 61 |  |
|  | Djoungolo | 19 (31.7) | 1 (1.7) | 40 (66.7) | 60 |  |
|  | Efoulan | 29 (44.6) | 0 (0.0) | 36 (55.4) | 65 |  |
|  | Mvog-Ada | 23 (44.2) | 0 (0.0) | 29 (55.8) | 52 |  |
|  | Nkolndongo | 19 (36.5) | 0 (0.0) | 33 (63.5) | 52 |  |
|  | Odza | 23 (46.0) | 0 (0.0) | 27 (54.0) | 50 |  |
| **Total** |  | 172 (42.4) | 1 (0.2) | 233 (57.4) | 406 |  |
| Cholera |  |  |  |  |  | 0.165 |
|  | Biyem-Assi | 2 (3.0) | 9 (13.6) | 55 (83.3) | 66 |  |
|  | Cite-Verte | 4 (6.6) | 8 (13.1) | 49 (80.3) | 61 |  |
|  | Djoungolo | 4 (6.7) | 7 (11.7) | 49 (81.7) | 60 |  |
|  | Efoulan | 2 (3.1) | 10 (15.4) | 53 (81.5) | 65 |  |
|  | Mvog-Ada | 0 (0.0) | 9 (17.3) | 43 (82.7) | 52 |  |
|  | Nkolndongo | 6 (11.5) | 5 (9.6) | 41 (78.8) | 52 |  |
|  | Odza | 1 (2.0) | 13 (26.0) | 36 (72.0) | 50 |  |
| **Total** |  | 19 (4.7) | 61 (15.0) | 326 (80.3) | 406 |  |

\* Fisher exact probability test

Biyem-Assi, Odza and Efoulan DHs had the highest number of HCWs fully vaccinated against COVID-19 (53%,46% and 45% respectively). Meanwhile, Nkoldongo, Djoungolo and Cite vert DHs had the highest percentage of HCWs fully vaccinated against Cholera (12%, 7% and 7% respectively) (**Figure 1**).

**Fig. 1.**
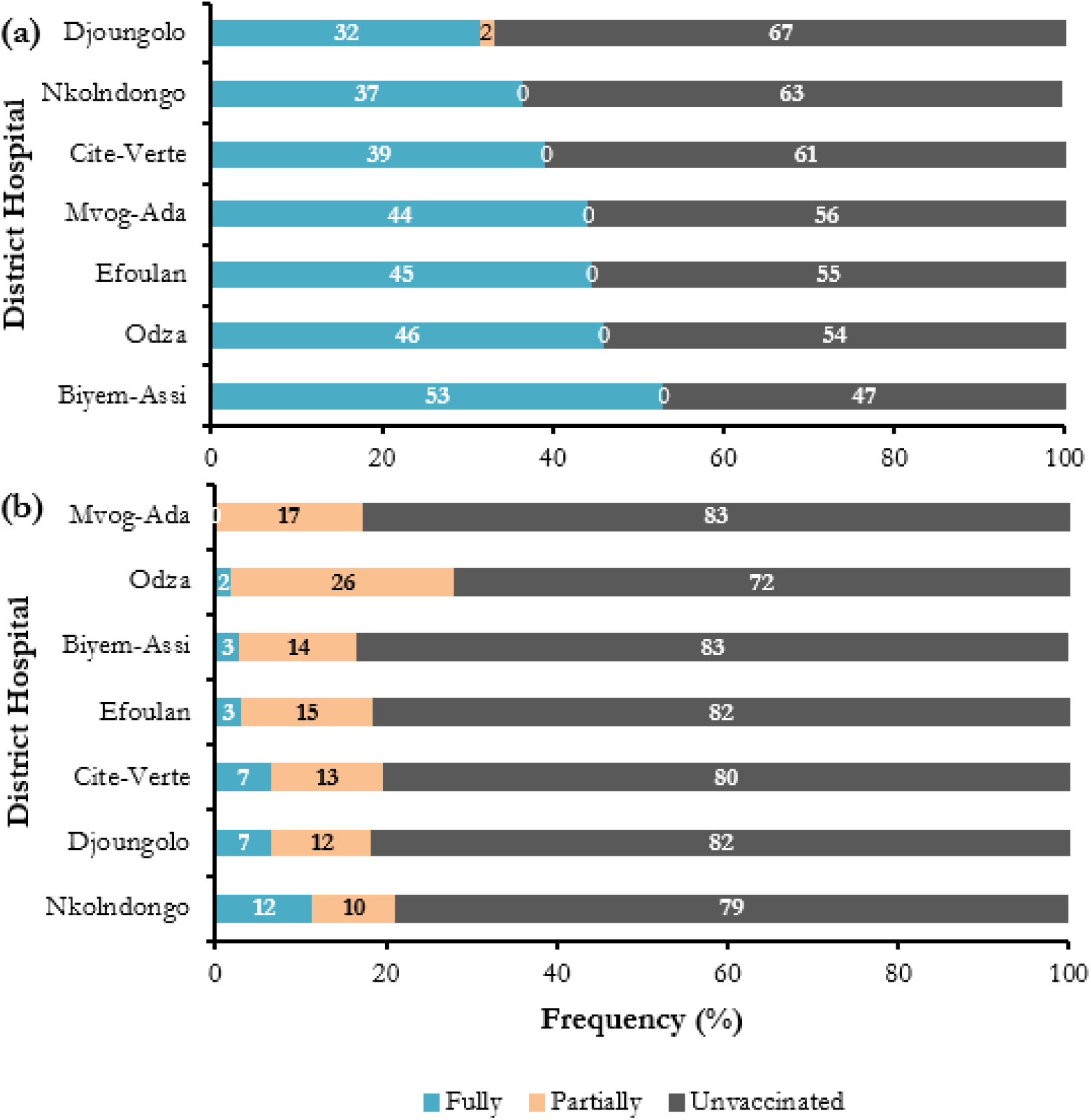
Vaccine coverage among health care workers in Yaoundé District Hospital, June 2024 (n=406) (a: COVID-19; b: Cholera)

### Predictors of COVID-19 vaccine uptake among HCWs

Univariate analysis showed that being female (*p*-value = 0.005), being a nurse (p-value < 0.001), laboratory technician (*p*-value = 0.011) and hygiene staff (*p*-value = 0.010) were predictors for COVID-19 update among HCWs (**Table 3**).

**Table 3.** Univariate analysis of factors associated with compliance to COVID-19 vaccination among HCWs in Yaoundé District Hospitals, June 2024 (n = 406)

| Predictor | Vaccination<br>n (%) |  | cOR | 95% CI |  | p-value |
| --- | --- | --- | --- | --- | --- | --- |
|  | Yes<br>n = 172 | No<br>n = 234 |  | Lower | Upper |  |
| **Sex** |  |  |  |  |  |  |
| Male | 54 (54.5) | 45 (45.5) | 1 |  |  |  |
| female | 118<br>(38.4) | 189<br>(61.6) | 1.92 | 1.22 | 3.05 | 0.005* |
| **Age group** |  |  |  |  |  |  |
| 19-29 | 14 (25.5) | 41 (74.5) | 1 |  |  |  |
| 30-44 | 111<br>(41.7) | 155 (58.3) | 0.48 | 0.24 | 0.90 | 0.026 |
| 45-54 | 42 (56.8) | 32 (43.2) | 0.26 | 0.12 | 0.55 | 0.001 |
| 55-66 | 5 (45.5) | 6 (54.5) | 0.41 | 0.11 | 1.62 | 0.190 |
| **Profession experience (in years)** |  |  |  |  |  |  |
| 1-4 | 29 (25.0) | 87 (75.0) | 1 |  |  |  |
| 5-14 | 106<br>(47.3) | 118<br>(52.7) | 0.37 | 0.22 | 0.60 | < 0.001 |
| 15-34 | 37 (56.1) | 29 (43.9) | 0.26 | 0.14 | 0.49 | < 0.001 |
| **Department** |  |  |  |  |  |  |
| Outpatient (vaccination) | 12 (40.0) | 18 (60.0) | 1 |  |  |  |
| Dentistry | 6 (33.3) | 12 (66.7) | 1.33 | 0.40 | 4.73 | 0.645 |
| Hygiene | 3 (20.0) | 12 (80.0) | 2.67 | 0.67 | 13.5 | 0.188 |
| Laboratory | 20 (41.7) | 28 (58.3) | 0.93 | 0.36 | 2.36 | 0.884 |
| Medicine | 33 (52.4) | 30 (47.6) | 0.61 | 0.25 | 1.45 | 0.266 |
| Gynecology | 17 (31.5) | 37 (68.5) | 1.45 | 0.57 | 3.68 | 0.432 |
| Pediatrics | 16 (40.0) | 24 (60.0) | 1.00 | 0.38 | 2.63 | > 0.999 |
| Surgery | 24 (52.2) | 22 (47.8) | 0.61 | 0.24 | 1.54 | 0.300 |
| Others | 41 (44.6) | 51 (55.4) | 0.83 | 0.35 | 1.90 | 0.662 |
| **Professional grade** |  |  |  |  |  |  |
| Doctor | 43 (62.3) | 26 (37.7) | 1 |  |  |  |
| Medical Student | 1 (25.0) | 3 (75.0) | 4.96 | 0.60 | 103 | 0.175 |
| Laboratory Technician | 21 (38.9) | 33 (61.1) | 2.60 | 1.26 | 5.48 | 0.011* |
| Nurse | 95 (38.2) | 154<br>(61.8) | 2.68 | 1.56 | 4.69 | < 0.001* |
| Hygiene staff | 3 (21.4) | 11 (78.6) | 6.06 | 1.71 | 28.7 | 0.010* |
| Other technician | 7 (58.3) | 5 (41.7) | 1.18 | 0.32 | 4.09 | 0.793 |
| Other staff | 2 (50.0) | 2 (50.0) | 1.65 | 0.19 | 14.5 | 0.625 |

cOR = Crude odds ratio; CI= Confidence Interval; \* $p$ -value □ 0.05

Multivariate analysis found that being a nurse (p-value = 0.001) and being a Laboratory Technician (p-value = 0.032) to be predictors of COVID-19 vaccine update among HCWs in Yaoundé (**Table 4**).

**Table 4.** Multivariate analysis of factors associated with compliance to COVID-19 vaccination among HCWs in Yaoundé District Hospitals, June 2024 (n = 406)

| Predictor | aOR | 95% CI limits |  | p-value |
| --- | --- | --- | --- | --- |
|  |  | Lower | Upper |  |
| Age group (in years) |  |  |  |  |
| 19-29 | 1 |  |  |  |
| 30-44 | 0.57 | 0.23 | 1.40 | 0.225 |
| 45-54 | 0.40 | 0.12 | 1.24 | 0.113 |
| 55-66 | 0.54 | 0.10 | 3.11 | 0.490 |
| Sex |  |  |  |  |
| Male | 1 |  |  |  |
| Female | 1.40 | 0.82 | 2.40 | 0.215 |
| Professional experience (in years) |  |  |  |  |
| 1-4 | 1 |  |  |  |
| 5-14 | 0.41 | 0.20 | 0.79 | 0.010 |
| 15-34 | 0.35 | 0.13 | 0.95 | 0.040 |
| Department |  |  |  |  |
| Outpatient (vaccination) | 1 |  |  |  |
| Dentistry | 2.30 | 0.59 | 9.48 | 0.234 |
| Laboratory | 0.59 | 0.06 | 3.92 | 0.604 |
| Internal Medicine | 0.82 | 0.30 | 2.17 | 0.693 |
| Gynecology | 1.63 | 0.59 | 4.48 | 0.343 |
| Pediatrics | 1.31 | 0.46 | 3.70 | 0.609 |
| Surgery | 0.76 | 0.27 | 2.14 | 0.608 |
| Others | 1.36 | 0.52 | 3.49 | 0.519 |
| Professional grade |  |  |  |  |
| Doctor | 1 |  |  |  |
| Medical Students | 2.06 | 0.21 | 45.9 | 0.561 |
| Nurses | 3.96 | 2.07 | 7.81 | 0.001* |
| Laboratory Technician | 8.00 | 1.38 | 69.8 | 0.032* |
| Other staff | 3.99 | 0.43 | 89.0 | 0.264 |

aOR = Adjusted odds ratio; CI = Confidence interval; \* $p$ -value □ 0.05

### Predictors of Cholera vaccine uptake among HCWs

Univariate analysis showed that working in the medical unit was a predictor associated with Cholera vaccine uptake among HCWs in our study (**Table 5**).

**Table 5.** Univariate analysis of factors associated with compliance to Cholera vaccination among HCWs in Yaoundé District Hospitals, June 2024 (n = 406)

| Predictor | Vaccination<br>n (%) |  | cOR | 95% CI |  | p-value |
| --- | --- | --- | --- | --- | --- | --- |
|  | Yes<br>n = 19 | No<br>n = 387 |  | Lower | Upper |  |
| **Age group (in years)** |  |  |  |  |  |  |
| 19-29 | 2 (3.6) | 53(96.4) | 1 |  |  |  |
| 30-44 | 8 (3.0) | 258(97.0) | 1.22 | 0.18 | 5.02 | 0.807 |
| 45-54 | 9(12.2) | 65 (87.8) | 0.27 | 0.04 | 1.11 | 0.106 |
| 55-66 | 0(0.0) | 11 (100.0) | NA |  |  |  |
| **Sex** |  |  |  |  |  |  |
| Male | 5 (5.1) | 94 (94.9) | 1 |  |  |  |
| female | 14 (4.6) | 293 (95.4) | 1.11 | 0.35 | 3.00 | 0.841 |
| **Professional experience (in years)** |  |  |  |  |  |  |
| 1-4 | 5 (4.3) | 111 (95.7) | 1 |  |  |  |
| 5-14 | 7 (3.1) | 217 (96.4) | 1.40 | 0.41 | 4.47 | 0.576 |
| 15-34 | 7 (10.6) | 59(89.4) | 0.38 | 0.11 | 1.24 | 0.111 |
| **Department** |  |  |  |  |  |  |
| Outpatient (vaccination) | 4 (13.3) | 26 (86.7) | 1 |  |  |  |
| Dentistry | 1 (5.6) | 17 (94.4) | 2.62 | 0.35 | 53.5 | 0.408 |
| Hygiene | 0 (0.0) | 15 (100.0) | NA |  |  |  |
| Laboratory | 2 (4.2) | 46 (95.8) | 3.54 | 0.65 | 26.8 | 0.160 |
| Medicine | 1 (1.6) | 62(98.4) | 9.54 | 1.33 | 191 | 0.048* |
| Gynecology | 3 (5.6) | 51 (94.4) | 2.62 | 0.54 | 14.1 | 0.230 |
| Pediatrics | 0(0.0) | 40 (100.0) | NA |  |  |  |
| Surgery | 3 (52.2) | 43 (93.5) | 2.21 | 0.45 | 11.9 | 0.325 |
| Others | 5 (5.4) | 87 (94.6) | 2.68 | 0.62 | 10.8 | 0.164 |
| **Professional grade** |  |  |  |  |  |  |
| Other technician | 1 (8.3) | 11 (91.7) | 1 |  |  |  |
| Laboratory technician | 2 (3.7) | 52 (96.3) | 2.36 | 0.10 | 26.9 | 0.498 |
| hygiene staff | 0 (0.0) | 14 (100.0) | NA |  |  |  |
| Doctor | 2 (2.9) | 67 (97.1) | 3.05 | 0.13 | 34.5 | 0.380 |
| Medical student | 0 (0.0) | 4 (100.0) | NA |  |  |  |
| Nurse | 14 (5.6) | 235 (94.4) | 1.53 | 0.08 | 8.74 | 0.696 |
| Other staff | 0 (0.0) | 4 (100.0) | NA |  |  |  |

aOR = Adjusted odds ratio, CI = Confidence interval, \*p-value < 0.05; NA = Not applicable

Working in the internal medicine ward (*p*-value = 0.046) and being a nurse (*p*-value = 0.001) were found to be predictors of Cholera vaccine uptake among HCWs in our study using multivariate analysis (**Table 6**).

**Table 6.**
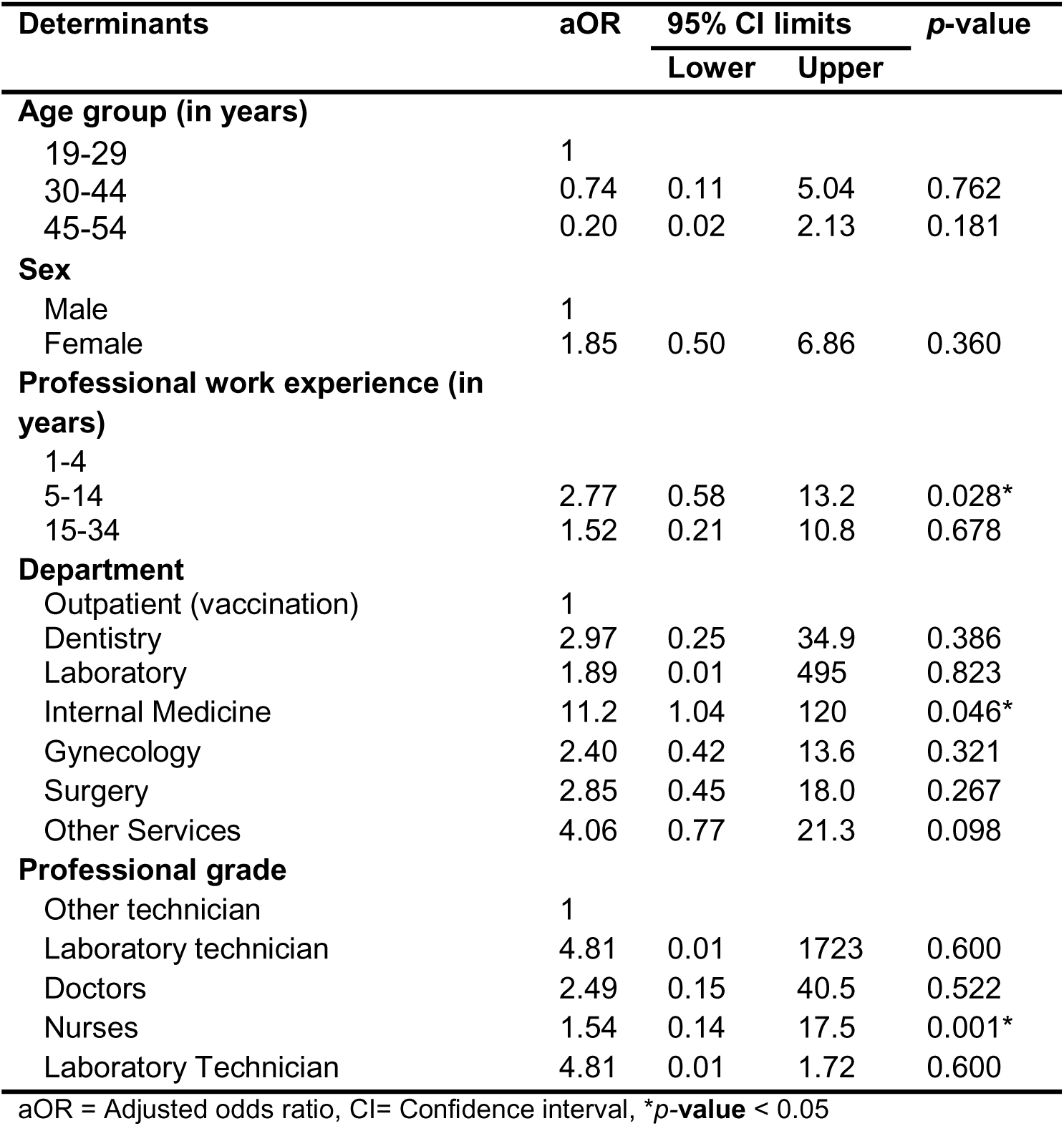
Multivariate analysis of factors associated with compliance to Cholera vaccination among HCWs in Yaoundé District Hospitals, June 2024 (n = 406)

### Knowledge on recommended vaccines for HCWs

Health care workers in our study reported hepatitis B (87.5%), COVID-19 (61.3%), tetanus (56.9%), cholera (18.7%) and meningitis vaccines as the top 5 vaccines recommended for them by guidelines (**Figure 2**).

**Fig. 2.**
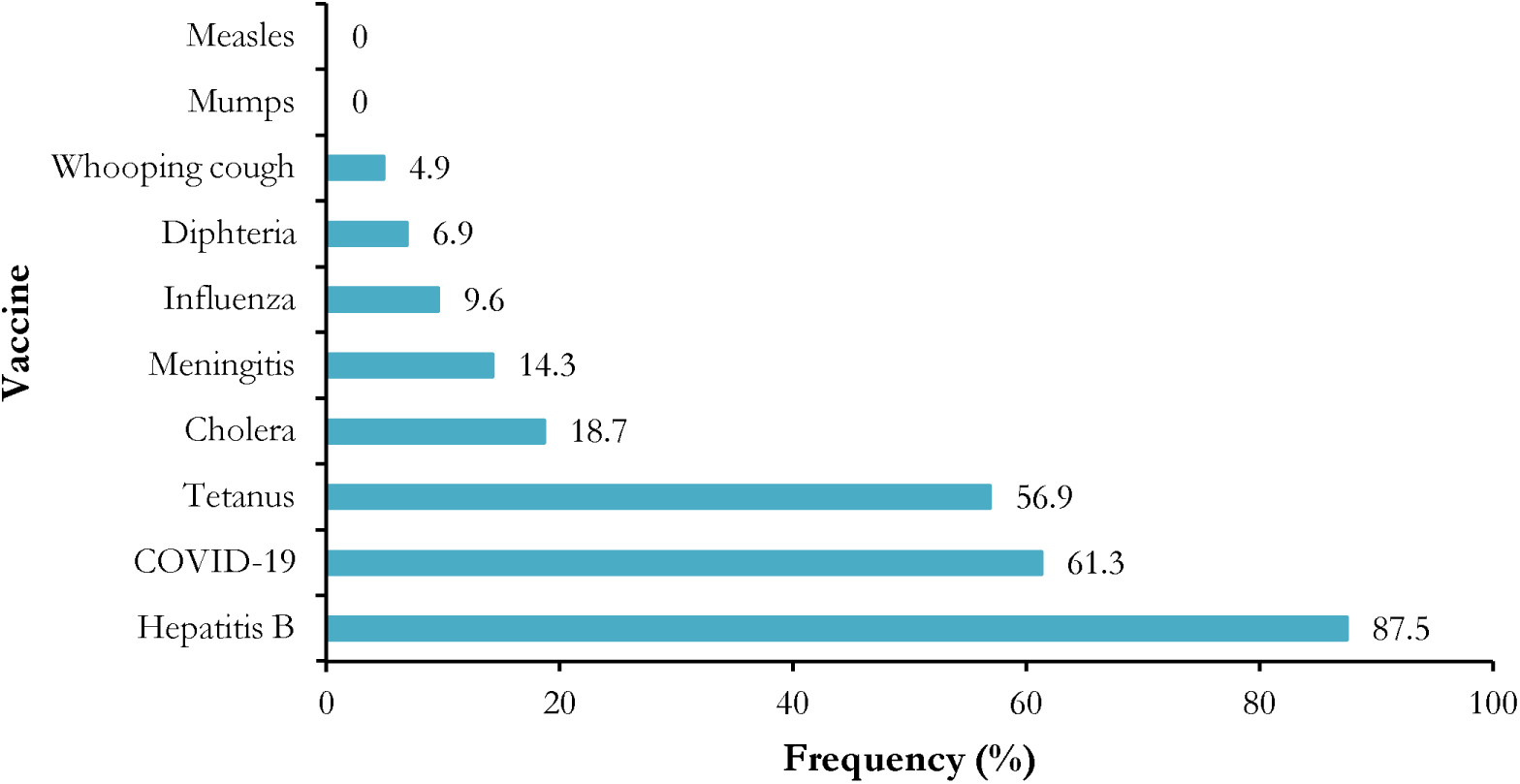
Knowledge of recommended vaccines according to international guidelines among healthcare workers in Yaoundé DHs (n = 406)

### Perceived risk of the working environment

Most of the HCWs (62.8%) perceived that their working environment was high risk for infection (Fig. 3).

**Fig. 3.**
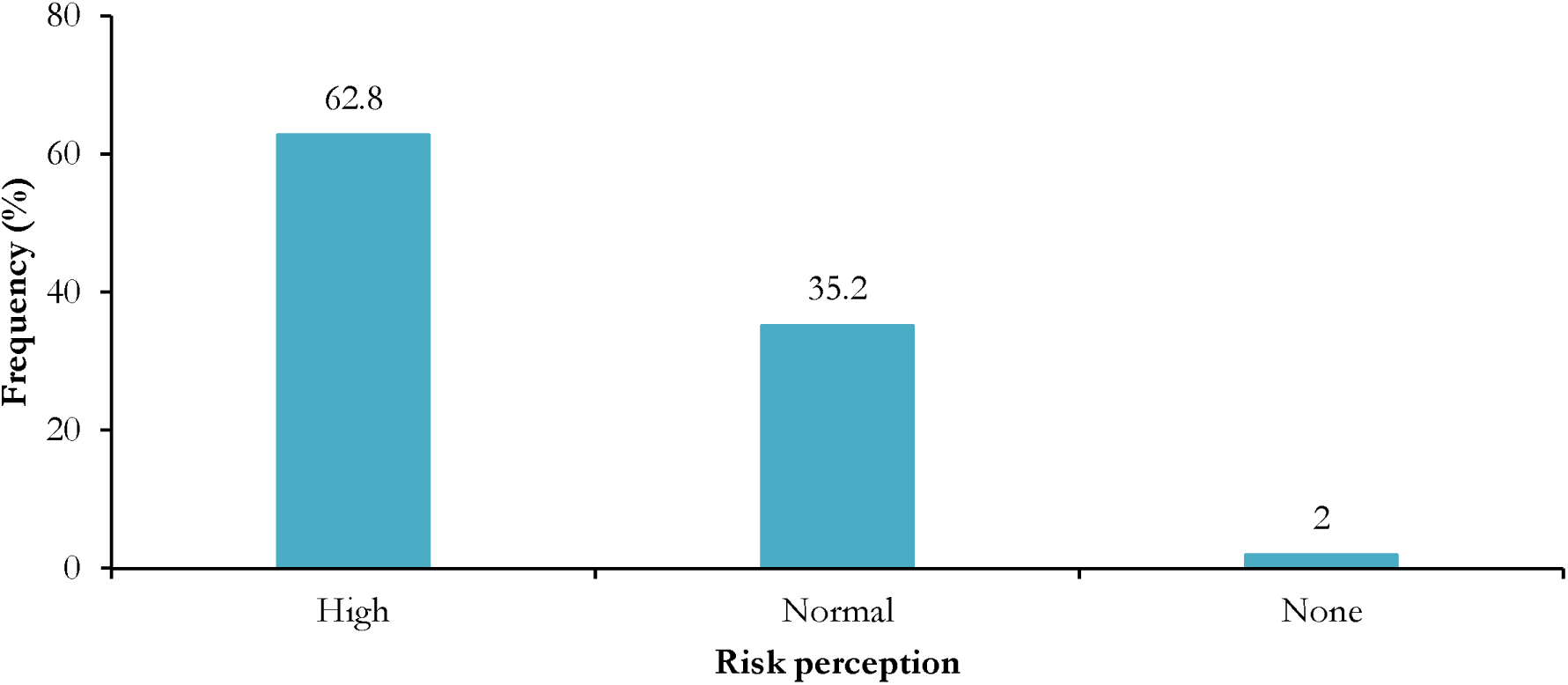
Risk perception of the working environment among HCWs in Yaoundé DHs

## Discussion

This study aimed to assess vaccine uptake and identify determinants of vaccination compliance among healthcare workers in Yaoundé. The results show that COVID-19 and cholera vaccination coverage was insufficient among healthcare workers, despite most being aware that they work in a high-risk environment.

The majority of participants were female and were nurse’s profession. Similar findings were reported in a study conducted in Nepal [24]. This predominance of women and nurses may be explained by the historical and social construction of the nursing profession, which has long been associated with caregiving qualities such as empathy, compassion, patience, and gentleness traits traditionally perceived as feminine. furthermore, nurses constitute the largest proportion of the healthcare workforce and play a crucial role in emergency management and in strengthening the resilience of health systems, which requires a substantial number of professionals in this category [25].

More than half of the participants had a university level education and between 5 and 14 years of professional experience. This may be explained by the increased recruitment in the public sector, particularly among healthcare workers, over the past fifteen years [26, 27]. Furthermore, The predominance of university level education observed in this study could also be related to the training requirements of health professions, which generally require at least a high school diploma, with approximately three years of training for nurses and laboratory technicians and about seven years for physicians [28, 29].

The proportion of healthcare workers who received the full doses of the COVID-19 vaccine in this study was 42.4%, which is considerably lower than the coverage reported in Finland (97.1 %) [30]. This difference may be explained by disparities in vaccine availability and distribution, differences in health system capacity, and unequal access to vaccination services between high-income and low- and middle-income settings [31]. Furthermore, vaccine hesitancy, concerns about vaccine safety and side effects, misinformation, and lower risk perception may have contributed to reduced vaccine uptake among healthcare workers in our context [32, 33]. These results underscore the need to strengthen communication strategies to build trust among healthcare workers in the COVID-19 vaccine’s reliability and the protection it offers.

Multivariate analysis revealed that being a nurse and a laboratory technician as significant predictors of COVID-19 vaccine uptake among healthcare workers. The higher likelihood of vaccination observed among nurses may be explained by their increased perception of infection risk due to frequent direct patient contact, as well as professional, ethical, and institutional expectations related to vaccination [34]. Similarly, laboratory technicians routinely handle biological specimens, including samples from patients infected with COVID-19, often prior to confirmed diagnosis, placing them at heightened occupational exposure. This frontline exposure may reinforce risk awareness and contribute to greater vaccine acceptance, primarily motivated by the desire to protect themselves and their families [35].

Less than 5% of healthcare workers were fully vaccinated against cholera, with limited awareness of the vaccine’s existence and concerns regarding its safety identified as major barriers to uptake [20]. Consequently, it is crucial to integrate behavior change communication sessions targeting healthcare workers to improve awareness and promote vaccine uptake [36].

Multivariate analysis identified working in the internal medicine ward and being a nurse as predictors of cholera vaccine uptake among healthcare workers. This finding is consistent with the literature, suggesting that nurses, as primary providers of direct patient care, often have higher risk perception and a stronger sense of professional responsibility, which may promote vaccine acceptance [37]. Additionally, internal medicine wards are frequently the first point of admission for undiagnosed patients, increasing occupational exposure to infections and potentially reinforcing adherence to preventive measures, including vaccination [38].

More than half of healthcare workers perceived their working environment as high-risk for infection, consistent with findings reported in Australia [39]. Occupational exposure to blood and other biological fluids has indeed been reported as high in hospital settings in Cameroon [40, 41]. This perception of risk, shared by the majority of healthcare professionals, represents an opportunity to strengthen targeted sensitization programs, improve risk communication, and integrate vaccination strategies into routine infection prevention and control policies for healthcare workers.

## Conclusion

This study highlights that although more than half of healthcare workers in Yaoundé were fully vaccinated against COVID-19, overall coverage remains insufficient to ensure optimal occupational protection. The very low uptake of the cholera vaccine emphasizes persistent gaps in comprehensive vaccination policies for HCWs in Cameroon. Demographic and professional characteristics including sex, age, professional category, work experience, and ward of assignment were significant determinants of vaccine uptake. To improve HCW immunization coverage, public health authorities should strengthen awareness and risk communication campaigns targeting younger and less experienced workers, integrate non-routine vaccines into occupational health programs, and ensure that essential vaccines are freely available within healthcare settings. Future research should explore qualitative aspects of vaccine hesitancy and institutional factors affecting vaccine access among HCWs. By addressing these barriers, Cameroon can enhance HCW protection, improve public trust in vaccination, and strengthen preparedness against both ongoing and emerging infectious disease threats.

## Data Availability

The datasets used and/or analyzed during the current study are available from the corresponding author on reasonable request.

## Declaration

### Ethics approval and consent to participate

The protocol was reviewed and approved by the Regional Human Health Committee of the Centre (CRERSH - Ce) ethical clearance: CE N° 2245/CRERSHC/2021 issued and from the Faculty of Medicine and Biomedical Sciences of the University of Yaoundé 1 IRB (N° 1115/UY1/FMSB/VDRC/DAASR/CSD). Informed consent was obtained prior inclusion. All study procedures were performed according to relevant guidelines of Helsinki Declarations.

### Consent for publication

Not applicable.

### Clinical trial number

Not applicable.

### Author’s contribution

Drafting of the study protocol, data collection, analysis and interpretation, drafting and editing of manuscript: A.N., F.Z.C.L; R.T. Critical revision of protocol, critical revision of manuscript: A.N., F.Z.C.L., F.N., I.T.; Conception, design and supervision of research protocol and implementation, data analysis plan, revision, editing and final validation of the manuscript: A.N., F.N., I.T.

## Acknowledgements

Our gratitude goes to all the healthcare workers who agreed to participate in this study and to the managers of the health facilities who gave their authorization for the conduct of this study.

